# “Knowledge practice gap of nurses towards COVID-19 patients’ dead body care in a tertiary care hospital”

**DOI:** 10.1101/2022.08.22.22278433

**Authors:** Girraj Saini, Mahendra Singh, Prasan Kumar Panda, Manish Kumar Sharma, Pankaj Punjot, Raviprakash Meshram, Puneet Kumar Gupta

## Abstract

**Aim:** To know the dead body care of COVID-19 patients.

**Objective:**

1. To determine health care professionals, knowledge, attitude, and practice towards Covid-19 dead body care.
2. To find the association of knowledge, attitude, practice with selected demographic variables.

**Background:** COVID-19 was a global pandemic and it was a serious note for health care professionals from many aspects. The virus was infective and causes serious infectionsto patients which were easily transmitted, hence specific dead body care is required for such kinds of patients. To keep this background in mind the study was conducted to identify the knowledge, practice and attitude towards COVID-19 dead body care among nurses.

**Methodology:** A cross sectional survey based study was done on 282 samples.Quantitative research design with purposive sampling technique data was collected for knowledge,attitude, and practice.

**Result:** Knowledge, attitude and practice were assessed and association was done with demographic profile. Hence the good knowledge, attitude and practicewere observed in experienced and trained nurses (p value<0.005. Whereas no significant changes were observed with age, gender and education qualification.

**Conclusion:** Overall knowledge, attitude and practice regarding COVID-19 dead body care were moderate to good. But it was important to identify the gap as it was a global pandemic and higher chances of spreading of infection.

## Introduction

The COVID pandemic is an ongoing global pandemic caused by SARS-COV-19, which is first identified during an outbreak in Wuhan, China. The 1^st^ case in India is reported in Thrissur district in Kerala followed by arising number of cases, as of the28^th^ of May 2022, there had been 43147530 confirmed cases and 524539 deaths (1–2). There are still some uncertainties in the natural history of COVID including source, transmissibility mechanisms, viral shedding and persistency of the virus in the environment. In health care settings, aerosol generating procedures, as well as the surfaces, fomites, and contaminated hands of healthcare workers (HCWs) have a role in the spread of disease. Since it is unclear how long the virus stays in the victim’s body, and the absence of data on infection from dead bodies, the WHO recommends the safe handling of bodies to prevent the possible spread of infection(ref). Therefore, the safety and well-being of persons handling the dead bodies should be the priority: it includes the use of personal protective equipment (PPE) while safe handling the bodies, placement of dead body in an impermeable bag to prevent leakage of body fluids before being moved from the isolation area or patient care room. (3–5)

For aerosol generating procedures for all confirmed or suspected COVID patients, all HCWs should follow the standard, contact, droplet and airborne precautions. All the persons including nursing staff handling the victims’ bodies in the isolation area, mortuary, ambulance, during crematorium, and burial ground should be trained in infection prevention and control practices. During hospital role of nursing staff remains crucial for dead body handling. Thus, this study aims to describe nurses knowledge, attitude, and practice gaps concerning COVID victim’s body care and factors that affect the above-mentioned precautions. So thatthe proper strategies to minimize the risk of infection in individuals handling the bodies of deceased persons infected with COVID are identified as the need of the hour. The information resulted from this study can serve as a vital tool to formulate relevant policies and guidelines during and after the outbreak and guide the nurses to prioritize self-protection and avoid occupational exposure.(6–8)

## Material and Methods

A single-centre cross-sectional survey-based study was conducted in the tertiary health care centre. To implement social distancing to avoid the spread of the COVID, it was not feasible to collect self-reported data-based surveys therefore the investigators used an online method of data collection. The sample size calculated by investigatorsas allhealth care workers posted in COVID units during data collection from July 2020 to September 2020. The survey commencedwith 282nurses posted in COVID units, and the required sample size was achieved on 20^th^ September 2020. All Nursing officers working in COVID units were recruited as study participants in the present study. A questionnaire was designed on Google forms, and a link was shared with whatsApp groups and personal messages of COVID unit staff.

### Measurement and Data Collection

The structured questionnaire was prepared after reviewing published literature based on WHO guidelines, institutional infection control protocols, and course material regarding emerging respiratory diseases including COVID(5–8). An initial draft of the questionnaire was designed and subsequently validated.The study instrument was sent to experts from the field of medicine, microbiology, nursing and toxicology,which was requested to give their expert opinion concerning its simplicity, relativity and importance. The questionnaire contains four parts socio-demographics, knowledge, attitude, and practice related items.

### Ethical considerations

Ethical clearance was obtained from theinstitute ethical committee vide letter no AIIMS/IEC/20/558 Dated 22/08/2020 before conducting the study. The study questionnaire contained a consent section that stated the purpose of the study, nature of the survey, study objectives, voluntary participation, declaration of confidentiality, and anonymity.

## Result

Response data of knowledge, attitude, and practices regarding COVID dead body care was collected from the 282 nurses. Demographic profile of study participants along with its association with knowledge is mentioned in Table 2. The majority of nurses were having work experience in COVID units. Whereas less the work experience shows significant low in knowledge (P=0.0055) for 0-30 days.

**Table 1:**
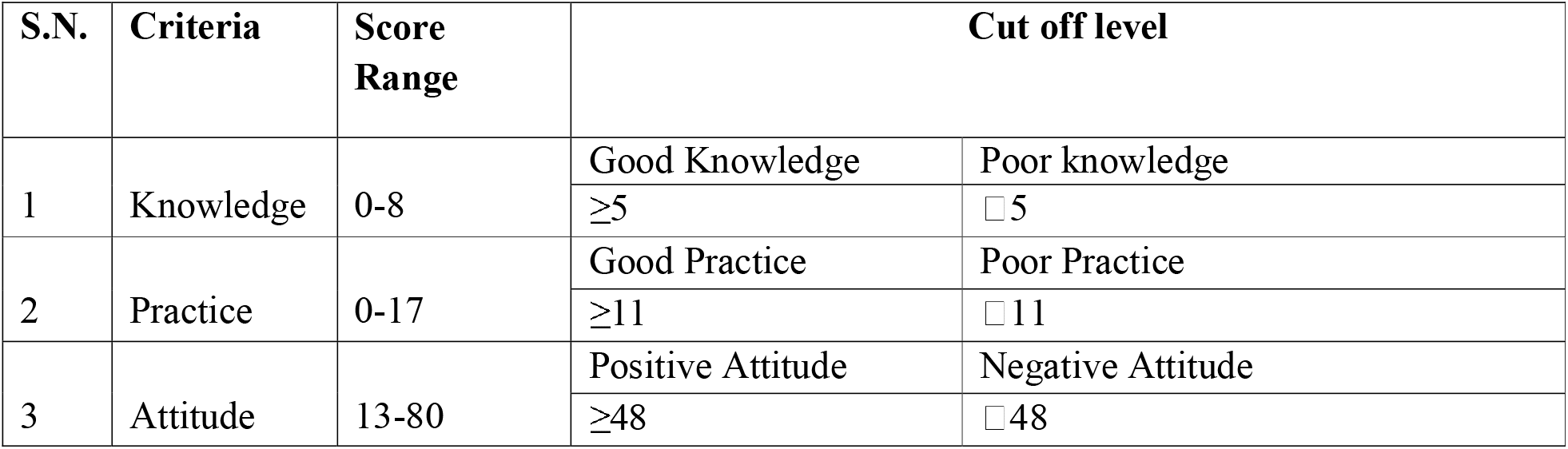
Criteria of score range with cut-off level.

| S.N. | Criteria | Score Range | Cut off level |  |
| --- | --- | --- | --- | --- |
| 1 | Knowledge | 0-8 | Good Knowledge | Poor knowledge |
|  |  |  | ≥5 | □ 5 |
| 2 | Practice | 0-17 | Good Practice | Poor Practice |
|  |  |  | ≥11 | □ 11 |
| 3 | Attitude | 13-80 | Positive Attitude | Negative Attitude |
|  |  |  | ≥48 | □ 48 |

**Table 2:** Association between demographic profile with Knowledge of Nurses (n=282)

|  |  |  |  |  |  |  |  |
| --- | --- | --- | --- | --- | --- | --- | --- |
| Variable | Good | % | Poor | % | Total | OR (95% CI) | p-Value |
| **Age** |  |  |  |  |  |  |  |
| 21-30 yr | 181 | 71.3 | 73 | 28.7 | 254 | 1 | 0.44 |
| >30 yr | 18 | 64.3 | 10 | 35.7 | 28 | 1.37(0.60-3.12) |  |
| **Gender** |  |  |  |  |  |  |  |
| Male | 137 | 71.7 | 54 | 28.3 | 191 | 1 |  |
| Female | 62 | 68.1 | 29 | 31.9 | 91 | 1.18(0.69-2.04) | 0.53 |
| **Years of experience in current profession** |  |  |  |  |  |  |  |
| <2 yr | 84 | 68.9 | 38 | 31.1 | 122 | 1.18(0.70-1.99) | 0.51 |
| 2-5 yr | 113 | 72.4 | 43 | 27.6 | 156 | 1 |  |
| >5 yr | 2 | 50 | 2 | 50 | 04 | 2.62(0.35-19.2) | 0.34 |
| **Work experience in COVID designated area** |  |  |  |  |  |  |  |
| 0-30 days | 24 | 52.2 | 22 | 47.8 | 46 | 2.65(1.33-5.27) | 0.0055 |
| 31-60 days | 48 | 73.8 | 17 | 26.2 | 65 | 1.02(0.52-1.99) | 0.94 |
| 61-80 days | 20 | 74.1 | 7 | 25.9 | 27 | 1.01(0.39-2.58) | 0.97 |
| >80 days | 107 | 74.3 | 37 | 25.7 | 144 | 1 |  |
| **Educational qualification** |  |  |  |  |  |  |  |
| General Nursing Midwifery | 28 | 58.3 | 20 | 41.7 | 48 | 1.93(1.01-3.68) | 0.43 |
| BSc/Msc Nursing | 171 |  | 63 |  | 234 | 1 |  |
| **Experience of handling dead body of COVID patient** |  |  |  |  |  |  |  |
| Yes | 151 | 83 | 31 | 17 | 182 | 1 |  |
| No | 48 | 48 | 52 | 52 | 100 | 5.27(3.04-9.15) | <0.001 |
| **Experience of handling dead body of SARS, MERSA, Swine Flu, NIPAH, EBOLA** |  |  |  |  |  |  |  |
| Yes | 104 | 86.7 | 16 | 13.3 | 120 | 1 |  |
| No | 95 | 58.6 | 67 | 41.4 | 162 | 4.58(2.48-8.45) | <0.001 |
| **Any training** |  |  |  |  |  |  |  |
| Yes | 119 | 85 | 21 | 15 | 140 | 1 |  |
| No | 80 | 56.3 | 62 | 43.7 | 142 | 4.39(2.48-7.76) | <0.001 |

The attitude gap was more significant in work experience in COVID designated area, educational qualification, the experience of handling dead bodies of COVID patients, and handling of other infections including the training (P<0.05) (Table 3). While age, gender, and experience of the current profession were not showing any differences in attitude.

**Table 3:** Association between demographic profile with Attitudeof Nurses (n=282)

|  |  |  |  |  |  |  |  |
| --- | --- | --- | --- | --- | --- | --- | --- |
| **Variable** | **Positive** | **%** | **Negative** | **%** | **Total** | **OR (95% CI)** | **p-Value** |
| **Age** |  |  |  |  |  |  |  |
| 21-30 yr | 80 | 31.5 | 174 | 68.5 | 254 | 1.88(0.85-4.14) | 0.115 |
| >30 yr | 13 | 46.4 | 15 | 53.6 | 28 | 1 |  |
| **Gender** |  |  |  |  |  |  |  |
| Male | 70 | 36.6 | 121 | 63.4 | 191 | 1 |  |
| Female | 23 | 25.3 | 68 | 74.7 | 91 | 1.71(0.98-2.98) | 0.058 |
| **Years of experience in current profession** |  |  |  |  |  |  |  |
| <2 yr | 33 | 27 | 89 | 73 | 122 | 8.09(0.81-80.5) | 0.074 |
| 2-5 yr | 57 | 36.5 | 99 | 63.5 | 156 | 5.21(0.52-51.2) | 0.157 |
| >5 yr | 3 | 75 | 1 | 4 | 4 | 1 |  |
| **Work experience in COVID designated area** |  |  |  |  |  |  |  |
| 0-30 days | 18 | 39.1 | 28 | 60.9 | 46 | 1 |  |
| 31-60 days | 19 | 29.2 | 46 | 70.8 | 65 | 1.55(0.70-3.45) | 0.277 |
| 61-80 days | 10 | 37 | 17 | 63 | 27 | 1.09(0.41-2.91) | 0.859 |
| >80 days | 46 | 31.9 | 98 | 68.1 | 144 | 0.067(0.03-0.14) | <0.0001 |
| **Educational qualification** |  |  |  |  |  |  |  |
| General Nursing Midwifery | 23 | 47.9 | 25 | 52.1 | 48 | 1 |  |
| BSc Nursing | 70 |  | 164 |  | 234 | 2.155(1.14-4.05) | 0.017 |
| **Experience of handling dead body of COVID patient** |  |  |  |  |  |  |  |
| Yes | 46 | 25.3 | 136 | 74.7 | 182 | 2.62(1.56-4.39) | 0.0002 |
| No | 47 | 47 | 53 | 53 | 100 | 1 |  |
| **Experience of handling dead body of SARS, MERSA, Swine Flu, NIPAH, EBOLA** |  |  |  |  |  |  |  |
| Yes | 22 | 18.3 | 98 | 81.7 | 120 | 3.47(1.99-6.06) | <0.0001 |
| No | 71 | 43.8 | 91 | 56.2 | 162 | 1 |  |
| **Any training** |  |  |  |  |  |  |  |
| Yes | 36 | 25.7 | 104 | 74.3 | 140 | 1.93(1.16-3.21) | 0.010 |
| No | 57 | 40.1 | 85 | 59.9 | 142 | 1 |  |

The practice of male nurses were having good practice compared to females (P=0.009) (Table 4). Age,education, years of nursing experience, dedicated COVID training, and experience of handling other infections had no significant practice gaps in dead body care. Work experiences in theCOVID unit and experience of handling the COVID dead body had significant practice gaps like more experiences had good practices (P<0.005).

**Table 4:**
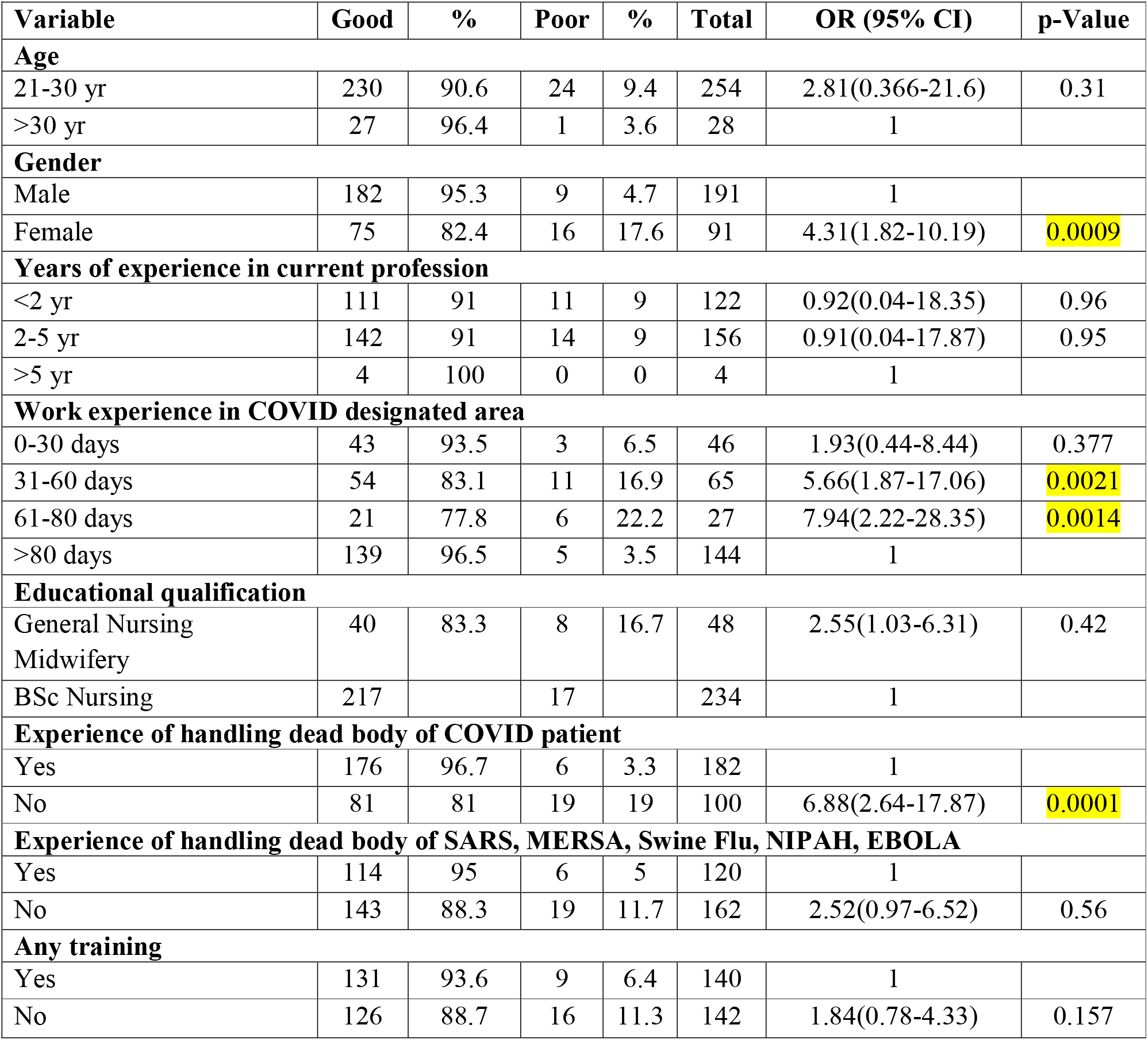
Association between demographic profile with Practice of Nurses (n=282)

## Discussion

The current study examined the knowledge, practice, and attitude of nurses regarding COVIDdead body care. Dead body care is an essential part of end-of-life care. The care after death especially during the major pandemic of COVIDhighlights many aspects for health care professionals to take care. In this study it is found that work experience in the COVID unit has a significant impact on knowledge-practice regarding COVID dead body care. Similarly, it is observed that training has improved the knowledge-practice of nursing personnel regarding dead body care.It is the first study that describes these observational findings on caring the COVID dead body by nurses.

The findings are strongly suggestive of good knowledge in those who worked in COVID areas. Many studies also suggested that effective and appropriate health education and training programme improve COVID knowledge and safe practices (8,9). COVID infection leads to many super-added infections and other health problems. Those who practice how to handle other infection like MERSA, SARS, NIPAH, etcare supposed to be well trained in handling a variety of infections (10).Similar findings were observed in thisstudy that those whohandles of other infection had higheraspects of knowledge, attitude, and practice.

It is well established fact that training improves knowledge, practice, and attitude(11,12). The same observations were observed among nurses while handling COVID dead body care except for practices. Nurses were more in exposure as they were direct in care providing to this global pandemic. Preparation for the response is a vital step tocope effectivelyand then deal with actual healthemergencies in the case of developing countries like India.Readiness for combating infectious diseases such as COVID begins with good understanding, positive thinking and safe and better practices. Similarly, in thestudy also attitude, and practice were assessed regarding handling of dead body care which gives direct exposure to the same(13,14).The dead body was considered infected and caring fora dead body with COVID infection for nurses requires a positive attitude with good knowledge and practical skills(15). The care during this war and specific protection with preventive aspects for the health care providers has a primary role in the reduction of the cases.

## Data Availability

All data produced in the present work are contained in the manuscript.

## Limitation

1. The study was conducted in one institute and can be conducted with different institutions.
2. There was no specific training provided regarding COVID-specific dead body care.
3. Study can be performed on a larger sample size.

## Conclusion

Overall knowledge, attitude, and practiceregarding COVID dead body care among nurses were moderate to good. As it ispandemic and has direct exposure to health care provider it was necessary to identify the gaps in the knowledge-practice of nurses regarding this dead body care.

